# Shifts in the pathogen spectrum and epidemiology of respiratory tract infections in the post-COVID-19 era: A study from Quzhou, Eastern China

**DOI:** 10.64898/2026.03.20.26348947

**Authors:** Ruijun Yang, Min Wang, Lei Lyu, Jialing You, Shiteng Huang, Bingdong Zhan

## Abstract

**Background:** Although the relaxation of COVID-19 containment measures in China has altered the transmission dynamics of respiratory pathogens, regional data on post-pandemic epidemiological characteristics remain limited.

**Objective:** This study aimed to investigate the pathogen spectrum and epidemiological characteristics of acute respiratory infections (ARIs) in Quzhou City from 2023 to 2024, providing a scientific basis for local prevention and control strategies.

**Methods:** A total of 2,800 respiratory specimens were collected from November 2023 to July 2024, comprising 1,960 influenza-like illness (ILI) cases from outpatient/emergency departments and 840 severe acute respiratory infection (SARI) cases from inpatient departments. All samples were tested for 13 common respiratory pathogens using multiplex fluorescence quantitative PCR. Etiological and epidemiological analyses were performed based on detection results and case information.

**Results:** The overall ARI positivity rate was 59.28% (1,660/2,800), with a male-to-female ratio of 1.07:1 (1,447/1,353). The three most prevalent pathogens were influenza virus (Flu, 23.21%, 650/2,800), Streptococcus pneumoniae (SP, 13.14%, 368/2,800), and adenovirus (ADV, 8.39%, 235/2,800). Single pathogen infections accounted for 73.55% (1,221/1,660) of positive cases, while co-infections with two or more pathogens accounted for 26.45% (439/1,660), yielding an overall co-infection rate of 15.68% (439/2,800). No significant gender difference was observed in detection rates. However, significant differences were found across case types, temporal periods, age groups, and geographic regions (P < 0.01). Children aged ≤5 years exhibited the highest positivity rate (78.00%, 378/525), while adults aged ≥65 years showed the lowest (34.53%, 144/417). Among surveillance regions, Kaihua County had the highest positivity rate (72.47%), and Changshan County the lowest (40.55%).

**Conclusions:** Multiple respiratory pathogens and co-infections are prevalent in Quzhou City, with distinct age-specific and seasonal patterns. These findings underscore the need for continuous multi-pathogen surveillance and integrated prevention strategies for influenza and other respiratory infectious diseases in the post-pandemic era.

## Introduction

Acute respiratory infections (ARIs) represent a significant global public health burden, ranking as the fourth leading cause of mortality worldwide [1]. The pathogen spectrum of respiratory tract infections is complex and dynamic, encompassing a wide array of etiological agents including viruses, bacteria, and atypical pathogens such as Mycoplasma pneumoniae and Chlamydia pneumoniae. These pathogens can circulate either independently or concomitantly, potentially leading to local outbreaks or even global pandemics [2].

In recent decades, the landscape of respiratory pathogens has been continually reshaped by the emergence of novel agents, including human metapneumovirus (HMPV) [3], human bocavirus (HBoV), and most notably, the unprecedented COVID-19 pandemic caused by SARS-CoV-2 [4]. The implementation of stringent non-pharmaceutical interventions during the pandemic dramatically altered the circulation patterns of common respiratory pathogens globally, leading to historically low detection rates of influenza, respiratory syncytial virus (RSV), and other endemic viruses [4]. This disruption of established epidemiological patterns has raised concerns about potential “immunity debt”—particularly among young children who experienced reduced exposure to common respiratory pathogens during the pandemic [5].

Following the relaxation of COVID-19 containment measures in China since early 2023, the country has witnessed a resurgence and alteration in the transmission dynamics of various respiratory pathogens. However, limited data are available regarding the post-pandemic epidemiological characteristics of respiratory pathogens at the regional level, particularly in medium-sized cities in Eastern China [6-8]. Understanding these patterns is crucial for optimizing diagnostic strategies, guiding empirical treatment, and implementing targeted prevention measures.

Quzhou City, located in western Zhejiang Province, has established a multi-pathogen surveillance system for acute respiratory infections. This study analyzed surveillance data from 2,800 ARI cases collected between November 2023 and July 2024, aiming to characterize the pathogen spectrum, epidemiological features, and age-specific distribution of respiratory pathogens in the post-COVID-19 era. Our findings will provide a scientific basis for the rapid and accurate diagnosis, prevention, and control of respiratory infectious diseases in the region.

## Materials and Methods

### 1. Ethics statement

This study was approved by the Ethics Committee of Quzhou Center for Disease Control and Prevention (Approval No.: IRB-2024-R-018, date of approval: 24 September 2024). Data were accessed for research purposes on 15/01/2025.The data used in this study were derived from the approved project titled “Research on infectious disease surveillance and early warning technology based on clinical syndromes and multi-pathogen detection techniques.”

Respiratory specimens used in this study were residual samples collected for routine clinical care purposes between November 2023 and July 2024. Due to the retrospective nature of the study and the use of fully anonymized residual specimens, the requirement for written informed consent was waived by the ethics committee. All patient information was anonymized prior to analysis to protect patient confidentiality.

The study was conducted in accordance with the principles of the Declaration of Helsinki and complied with the national regulations for biomedical research involving human subjects in China.

### 2. Materials

#### 2.1 Study Subjects and Data Collection

A total of 1,960 influenza-like illness (ILI) cases from outpatient and emergency departments, and 840 severe acute respiratory infections (SARI) cases from inpatient departments were enrolled from county/district people’s hospitals across Quzhou City between November 2023 and July 2024. Demographic and clinical information of the enrolled cases was recorded in detail. The inclusion criteria [9] were: (1) ILI: fever (body temperature ≥38°C) accompanied by either sore throat or cough. (2) SARI: patients presenting with the following clinical features upon admission or during hospitalization: history of fever (axillary temperature ≥38°C) at onset, accompanied by cough or sore throat, and illness onset within the last 10 days. The case enrollment principles complied with the “Notice on the issuance of the Zhejiang Province Acute Respiratory Infection Multi-pathogen Surveillance Protocol (Trial)” issued by the Comprehensive Office of the Zhejiang Provincial Disease Control and Prevention Bureau.

#### 2.2 Specimen Collection

1. For ILI cases: Throat swabs, nasal swabs, or nasopharyngeal swabs were collected within 3 days of illness onset. After collection, specimens were placed into collection tubes containing 3–4 mL of viral transport medium.
2. For SARI cases: Respiratory specimens, including throat swabs, nasal swabs, nasopharyngeal swabs, bronchoalveolar lavage fluid, and pleural puncture fluid, were collected from patients admitted to the hospital due to ARI, within 3 days of illness onset. After collection, specimens were placed into collection tubes containing 3–4 mL of viral transport medium.

#### 2.3 Instruments and Reagents

ABI real-time fluorescent PCR instruments and a ZY Biotechnology nucleic acid extraction system were used. PCR detection was performed using a 13-pathogen nucleic acid detection kit (Fluorescence PCR method) manufactured by Jiangsu Bioperfectus Technologies Co., Ltd.

### 3. Methods

Specimens were transported to the virology laboratory at 4°C immediately after collection. Real-time RT-PCR was performed promptly to detect nucleic acids of influenza virus (Flu), Mycoplasma pneumoniae (MP), Streptococcus pneumoniae (SP), SARS-CoV-2, rhinovirus (RV), human metapneumovirus (HMPV), human adenovirus (AdV), respiratory syncytial virus (RSV), human parainfluenza virus (HPIV), human coronavirus (HCoV), human bocavirus (HBoV), Chlamydia pneumoniae (CP), and enterovirus (EV).

### 4. Statistical Analysis

Data were collated using Excel and analyzed using SPSS 21.0 software. Comparisons of rates among multiple groups were performed using the chi-square test (χ^2^ test). A trend test was used to analyze the association between age and the detection rate of co-infections. A P-value <0.05 was considered statistically significant.

## Results

### 1 Baseline Characteristics

From November 2023 to July 2024, a total of 2,800 acute respiratory infection (ARI) surveillance cases were enrolled, including 1,960 ILI cases (70.0%) and 840 SARI cases (30.0%). Among these, there were 1,447 males(51.7%) and 1,353 females(48.3%), with a male-to-female ratio of 1.07:1. The age distribution was as follows: ≤5 years (n=525, 18.8%), 6–14 years (n=713, 25.5%), 15–64 years (n=1,145, 40.9%), and ≥ 65 years (n=417, 14.9%).

### 2 Etiological Characteristics

#### 2.1 Overall Pathogen Detection

Nucleic acid testing for 13 common respiratory pathogens was performed on all 2,800 enrolled cases. A total of 1,660 cases (59.28%, 1,660/2,800) tested positive for ARI pathogens. The detection rate in ILI cases (64.39%) was significantly higher than that in SARI cases (47.38%) (P < 0.01). No statistically significant difference in pathogen detection was observed between sexes (P = 0.545). However, significant differences in detection rates were found among different age groups (P < 0.01). The highest positivity rate was observed in the 0–5 years age group (78.00%, 378/525), while the lowest was in the >65 years age group (34.53%, 144/417). A significant linear trend was observed, indicating that the positivity rate decreased with increasing age (χ^2^ = 172.511, P < 0.01). Furthermore, pathogen detection rates varied significantly across different months (P < 0.01). The baseline characteristics of the monitored cases are presented in Table 1.

**Table 1.**
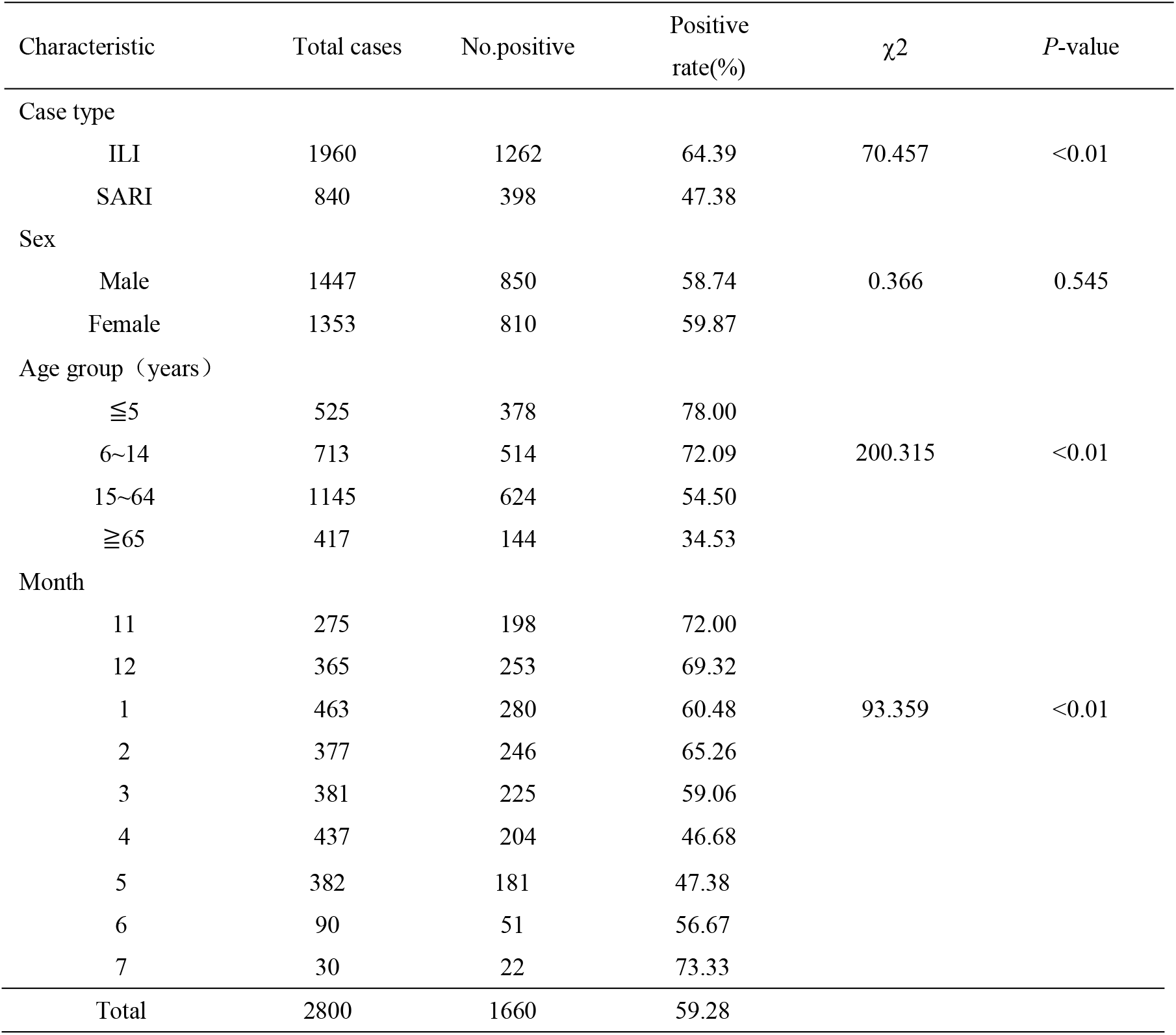
Baseline characteristics of acute respiratory infection surveillance cases in Quzhou City.

#### 2.2 Detection Rates of Specific Pathogens

Among the 2,800 cases, the detection rates of SARS-CoV-2, influenza virus (Flu), respiratory syncytial virus (RSV), human adenovirus (AdV), human metapneumovirus (HMPV), rhinovirus (RV), human parainfluenza virus (HPIV), human coronavirus (HCoV), human bocavirus (HBoV), enterovirus (EV), Mycoplasma pneumoniae (MP), Chlamydia pneumoniae (CP), and Streptococcus pneumoniae (SP) are shown in Table 2. Influenza virus exhibited the highest detection rate (23.21%), followed by Streptococcus pneumoniae (13.14%) and human adenovirus (8.39%), ranking second and third, respectively.

**Table 2.**
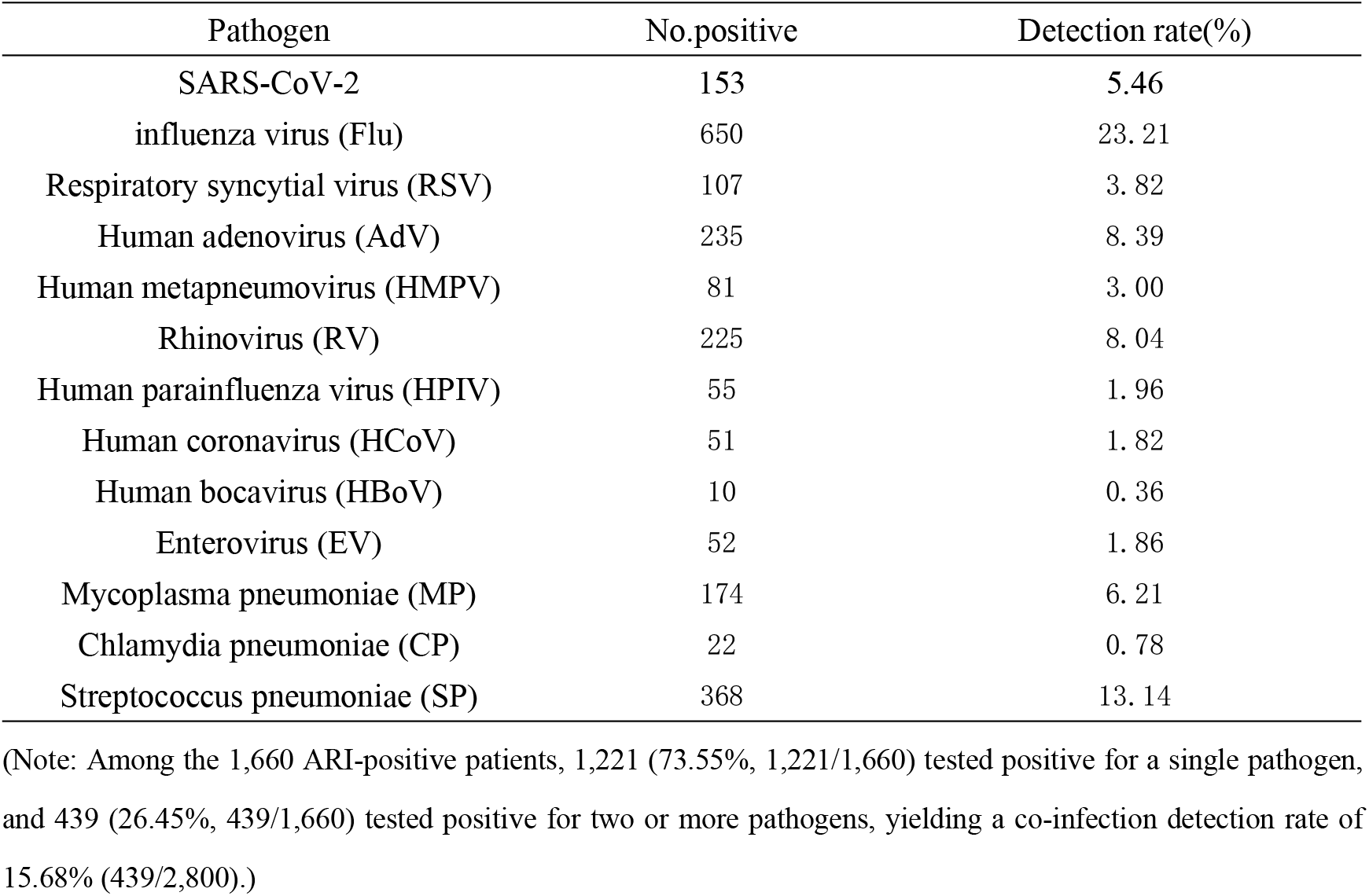
Detection of pathogens in acute respiratory infection surveillance cases in Quzhou City.

### 3. Epidemiological Characteristics

#### 3.1 Age Distribution

The difference in pathogen detection rates among different age groups was statistically significant (χ^2^ = 200.315, P < 0.01). The highest positivity rate was observed in children aged 5 years and below (78.00%, 378/525), while the lowest was in cases aged over 65 years (34.53%, 144/417). Among children aged 5 years and below, Streptococcus pneumoniae (SP) showed the highest detection rate (20.00%), followed by influenza virus (FLU) (15.24%) and adenovirus (ADV) (13.90%). In contrast, influenza virus (FLU) was the most frequently detected pathogen in the adolescent group (6–14 years), adult group (15–64 years), and elderly group (>65 years), with detection rates of 27.21%, 28.65%, and 11.51%, respectively. The differences in pathogen detection rates across all age groups were statistically significant (P < 0.05). Detailed pathogen detection results for different age groups are presented in Table 3.

**Table 3.**
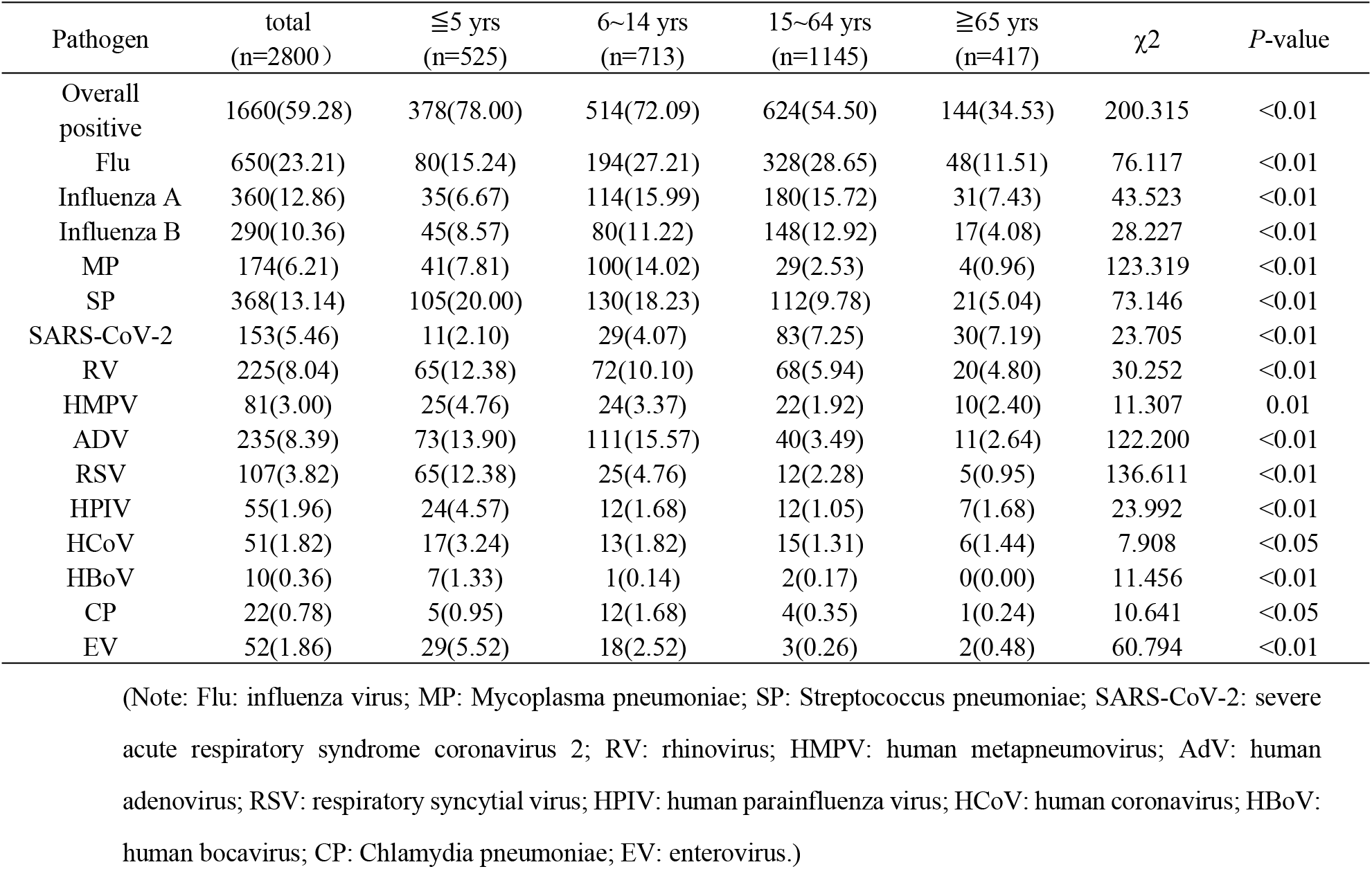
Pathogen detection in ARI cases by age group [n (%)]

#### 3.2 Time Distribution

From a temporal perspective, the monthly positive rate of pathogen detection throughout the surveillance period from November 2023 showed an overall trend of initially decreasing, then increasing, followed by another decrease, with inflection points occurring in February 2024 and April 2024. Specifically, the dominant pathogen, influenza A virus, exhibited peak detection rates in November (45.45%) and December (43.29%), subsequently returning to relatively stable levels. Influenza B virus showed peak detection rates in January (27.86%) and February (29.71%). Streptococcus pneumoniae (SP) was detected at relatively high levels throughout the entire study period. Adenovirus (AdV) showed a peak detection period from March to May. Additionally, SARS-CoV-2 exhibited a peak between February and March. In April, all 13 pathogens were detected. The temporal distribution characteristics of various pathogens detected in the 2,800 acute respiratory infection surveillance cases are presented in Table 4.

**Table 4.**
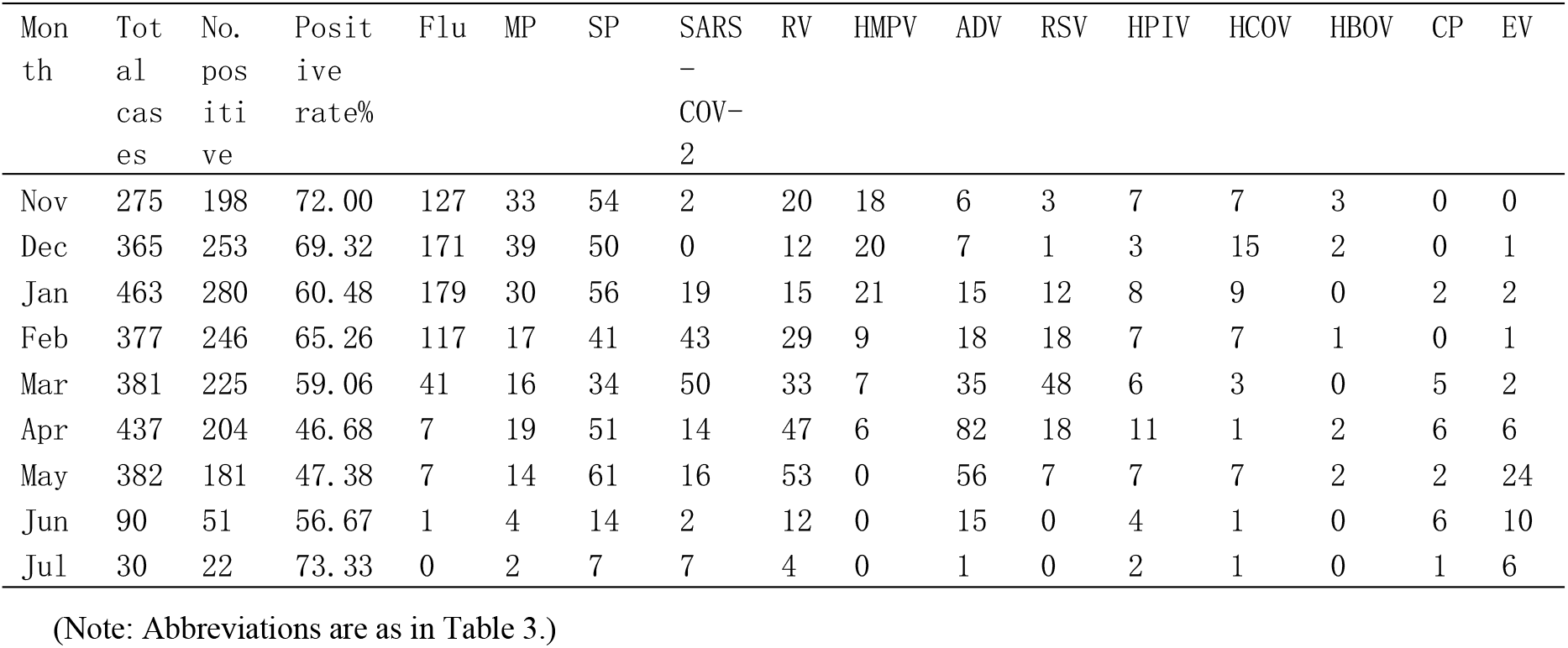
Temporal distribution of pathogen detection in ARI cases in Quzhou City.

#### 3.3 Geographic Distribution

Analysis of pathogen detection across different counties and districts revealed the following: In Kecheng District, 360 ARI cases tested positive (66.67%, 360/540), with the top three pathogens being Flu (25.74%, 139/540), SP (22.78%, 123/540), and RV (11.30%, 61/540). In Qujiang District, 261 ARI cases tested positive (57.49%, 261/454), with the top three pathogens being Flu (22.90%, 104/454), SP (12.56%, 57/454), and RV (10.79%, 49/454). In Longyou County, 361 ARI cases tested positive (67.10%, 361/538), with the top three pathogens being Flu (24.54%, 132/538), SP (13.57%, 73/538), and RV (10.78%, 58/538). In Jiangshan City, 171 ARI cases tested positive (45.60%, 171/375), with the top three pathogens being Flu (22.40%, 84/375), AdV (5.07%, 19/375), and SP (3.73%, 14/375). In Changshan County, 178 ARI cases tested positive (40.55%, 178/439), with the top three pathogens being Flu (18.91%, 83/439), SP (7.74%, 34/439), and SARS-CoV-2 (6.15%, 27/439). In Kaihua County, 329 ARI cases tested positive (72.47%, 329/454), with the top three pathogens being Flu (23.79%, 108/454), AdV (20.48%, 93/454), and SP (14.76%, 67/454).Notably, Kaihua County exhibited the highest positivity rate (72.47%), while Changshan County had the lowest (40.55%). Furthermore, the difference in respiratory multi-pathogen positivity rates among the surveillance regions was statistically significant (χ^2^ = 152.049, P < 0.01).

### 4 Comparison of Pathogen Detection between ILI and SARI Cases

Overall, the difference in pathogen detection rates between ILI and SARI cases was statistically significant (χ^2^ = 70.457, P < 0.01). Specifically, statistically significant differences (P < 0.05) between ILI and SARI cases were observed for Flu, SP, SARS-CoV-2, and RSV, while no significant differences were found for the remaining pathogens (P > 0.05).The top five pathogens detected in ILI cases were Flu (28.06%, 550/1960), SP (14.80%, 290/1960), RV (8.21%, 161/1960), SARS-CoV-2 (6.48%, 127/1960), and AdV (5.78%, 171/1960). The top five pathogens detected in SARI cases were Flu (11.90%, 100/840), SP (9.29%, 78/840), AdV and RV (tied for third, 7.62%, 64/840 each), MP (7.38%, 62/840), and RSV (5.12%, 43/840).The detection rates of Flu, SP, RV, HMPV, AdV, HCoV, EV, and SARS-CoV-2 were lower in SARI cases compared to ILI cases, whereas the detection rates of RSV, HPIV, HBoV, CP, and MP were higher in SARI cases than in ILI cases. Detailed comparisons are presented in Table 5.

**Table 5.**
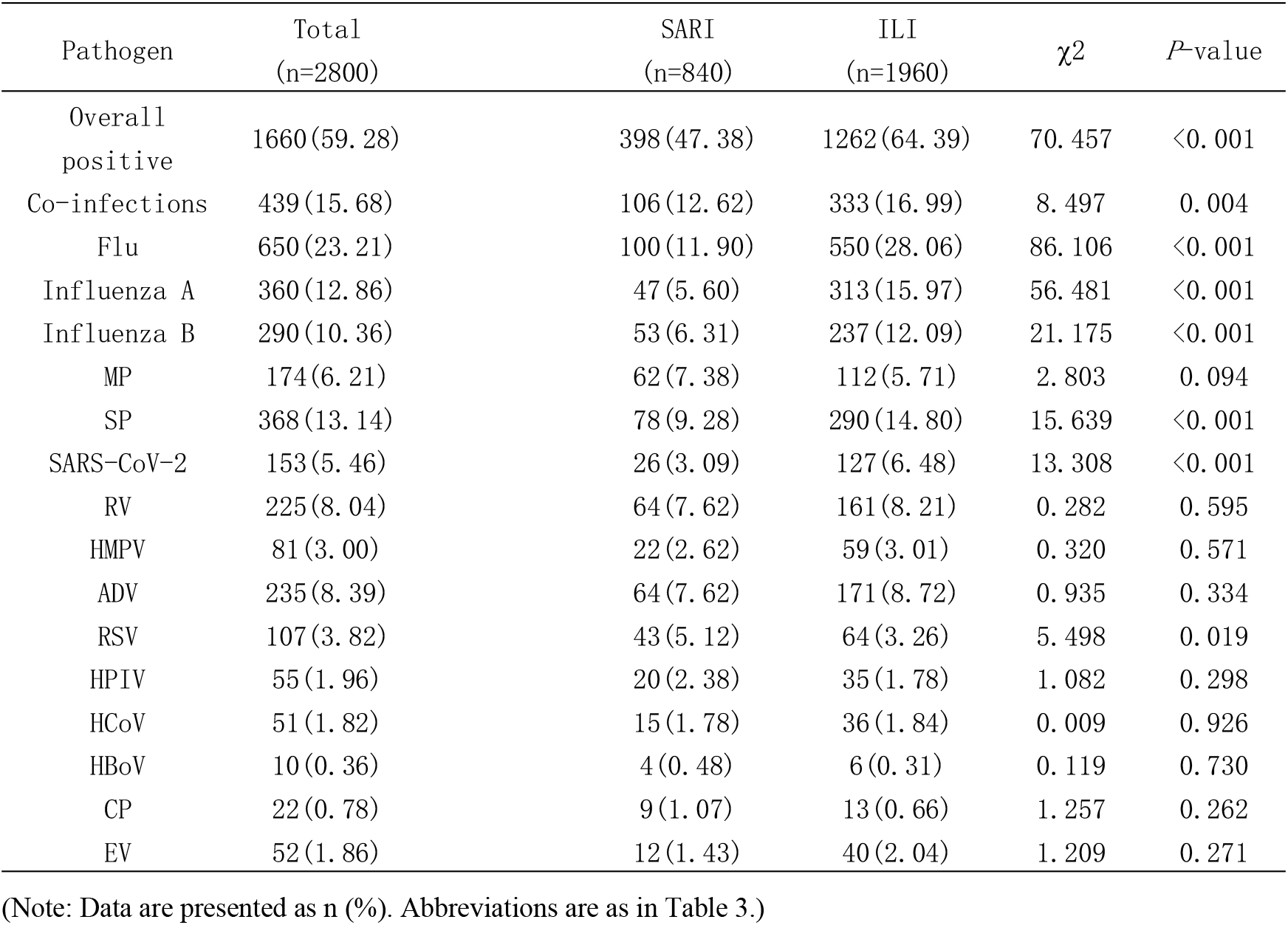
Comparison of pathogen detection rates between ILI and SARI cases [n (%)]

## Discussion

Acute respiratory infections (ARIs) are the fourth leading cause of death globally, imposing a substantial disease burden on public health worldwide[10]. The epidemiological patterns of respiratory pathogens have undergone significant changes in the context of the COVID-19 pandemic, with marked declines in the detection of common ARIs observed compared to other years [4]. The implementation of COVID-19 prevention and control measures reduced opportunities for exposure to respiratory pathogens, consequently leading to a diminished population-level immune barrier against common respiratory pathogens, particularly among young children. This immune naivety may have contributed to more severe circulation and transmission of respiratory pathogens following the conclusion of the pandemic [5]. The respiratory tract provides an ecological niche for the coexistence of multiple pathogens across temporal and spatial dimensions, potentially leading to interactions among different pathogens and resulting in changes to epidemiological patterns, including seasonality, activity intensity, and co-infection dynamics [2]. The year 2023 marked the first year in China when COVID-19 management was adjusted from Class A to Class B. Analysis of multi-pathogen surveillance data for acute respiratory infections in Quzhou City during 2023-2024 is conducive to understanding the epidemiological distribution characteristics of other respiratory pathogens in the Quzhou region after the COVID-19 pandemic, thereby providing a scientific basis for formulating subsequent respiratory infection prevention and control strategies.

The overall ARI positivity rate in our study (59.28%) was higher than that previously reported in Beijing (31.1%) [6] but lower than that reported in Ganzhou City in 2021 (61.11%) [7]. No statistically significant difference in pathogen positivity rates was observed between sexes (P =0.545). However, children aged 5 years and below exhibited the highest positivity rate, while individuals aged≥65 years showed the lowest positivity rate. This finding may be attributed to the implementation of a public health program in Quzhou City in recent years, providing free influenza vaccination to residents aged 60 years and above.During 2023-2024, the overall positivity rate of acute respiratory pathogens in Quzhou City showed a fluctuating trend, which is consistent with the seasonal epidemiological characteristics of respiratory infectious diseases. Beginning in November 2023, the monthly pathogen detection rate demonstrated an overall pattern of initial decrease, followed by an increase, and then another decrease, with inflection points occurring in February 2024 and April 2024. Furthermore, significant differences in respiratory multi-pathogen positivity rates were observed across surveillance regions, with Kaihua County showing the highest detection rate and Changshan County the lowest. This variation may be related to differences in the professional expertise of medical staff and sampling practices across different regions.Regarding pathogen detection rates, ILI cases showed significantly higher positivity rates compared to SARI cases. It is speculated that this may be related to the fact that infections caused by most respiratory pathogens included in this study typically present with mild symptoms and a relatively low proportion of severe cases, in addition to the likelihood that SARI cases had already received treatment prior to sampling.

The top three pathogens detected in this study were influenza virus (Flu), Streptococcus pneumoniae (SP), and adenovirus (AdV). Pathogen positivity was predominantly characterized by single infections, with an overall co-infection detection rate of 15.68%. This was higher than the co-infection rate of 10.80% reported in a study from Tongzhou District, Beijing [8], but fell within the range of 0.60% to 27.00% reported for respiratory pathogen co-infection rates in domestic and international studies [11-12]. In 2023, the WHO issued a statement indicating that Mycoplasma pneumoniae (MP) is one of the pathogens contributing to the increasing number of pediatric community-acquired pneumonia (CAP) cases in China [13]. Streptococcus pneumoniae (SP), as an important opportunistic pathogen, primarily colonizes the nasopharynx of children and can cause various diseases when immune function declines [14]. This may explain the relatively high detection rates of SP and MP observed in individuals under 14 years of age in this study.Human rhinovirus (RV) is one of the major pathogens causing the common cold. Rhinovirus infection typically presents as a mild, self-limiting illness, with most common manifestations being upper respiratory symptoms such as nasal congestion, rhinorrhea, and sore throat. However, it can cause severe disease in infants, the elderly, and immunocompromised individuals [15]. Surveillance data from this study showed that rhinovirus was detected throughout the entire study period, suggesting that continuous surveillance of rhinovirus should be maintained.Human parainfluenza virus (HPIV) is also an important respiratory pathogen that can cause bronchitis and pneumonia in infants and immunocompromised populations, ranking as the second most common pathogen causing acute respiratory infections in children after respiratory syncytial virus [16]. In Quzhou, children aged 0-5 years were the most susceptible population for both respiratory syncytial virus and parainfluenza virus infections.Human metapneumovirus (HMPV) is widely prevalent worldwide, with high infection and mortality rates [3,17]. In this surveillance, relatively higher detection rates of HMPV were observed in children aged 0-5 years (4.76%) and individuals aged 6-14 years (3.37%), indicating the need to strengthen surveillance for HMPV across all age groups.

Through multi-pathogen detection and analysis of ARI cases in Quzhou following the COVID-19 pandemic, this study preliminarily identified the circulation of multiple pathogen types and co-infections of respiratory pathogens in the region. The main limitations of this study include the limited number of laboratory-tested specimens, restriction to the 13 pathogens included in the investigation, and consequently limited representativeness. Additionally, some pathogens have long epidemic cycles, and short-term surveillance results are insufficient to reflect their complete epidemiological characteristics. Therefore, the pathogen spectrum composition of ARI cases cannot be fully elucidated, necessitating more systematic analysis of ARI pathogen profiles in Quzhou in the future. Although multi-pathogen surveillance for acute respiratory infections has been initiated in Quzhou, it remains in the preliminary stage, with insufficient research on refined genotyping, genetic variation, and co-infection patterns of various viruses and bacteria. Therefore, further improvement of the multi-pathogen surveillance system is needed, along with integration of relevant clinical data to analyze the causes of respiratory infections, thereby contributing to early detection, prevention, control, and treatment of acute respiratory infectious diseases.

## Acknowledgments

We extend our sincere gratitude to our colleagues from Quzhou Center for Disease Control and Prevention and the county-level Centers for Disease Control and Prevention (Kecheng, Qujiang, Longyou, Jiangshan, Changshan, Kaihua) for their invaluable support in sample collection and data management. We thank the clinical staff at all participating hospitals for their assistance with case enrollment. Most importantly, we thank the patients who agreed to participate in this study, without whom this research would not have been possible.

## Institutional Review Board Statement

The study was conducted in accordance with the Declaration of Helsinki and approved by the Ethics Committee of Quzhou Center for Disease Control and Prevention (Approval No.: IRB-2024-R-018, date of approval: 24 September 2024). The requirement for informed consent was waived for the use of retrospective anonymized residual clinical specimens collected prior to ethical approval.

## Informed Consent Statement

Written informed consent was obtained from all participants or their legal guardians for prospective sample collection. For the retrospective use of residual specimens collected between November 2023 and July 2024 (prior to ethical approval), the requirement for informed consent was waived by the ethics committee due to the retrospective nature of the study, the use of fully anonymized samples, and the minimal risk to participants.

## Data Availability Statement

Relevant data are within the manuscript. The datasets generated and analyzed during the current study are available from the corresponding author upon reasonable request.

## Author Contributions

Conceptualization: Ruijun Yang, Min Wang, Shiteng Huang, Bingdong Zhan; Methodology: Ruijun Yang, Lei Lyu, Bingdong Zhan; Formal analysis: Ruijun Yang, Min Wang; Investigation: Ruijun Yang, Min Wang, Jialing You; Resources: Ruijun Yang, Shiteng Huang;

Data curation: Ruijun Yang, Min Wang; Writing–original draft: Ruijun Yang, Jialing You;Writing–review & editing: Min Wang, Lei Lyu, Shiteng Huang; Visualization: Ruijun Yang, Bingdong Zhan; Supervision: Shiteng Huang, Bingdong Zhan; Project administration: Shiteng Huang, Bingdong Zhan; Funding acquisition: Shiteng Huang, Bingdong Zhan. All authors have read and agreed to the published version of the manuscript.

